# The Paxlovid Rebound Study: A Prospective Cohort Study to Evaluate Viral and Symptom Rebound Differences Between Paxlovid and Untreated COVID-19 Participants

**DOI:** 10.1101/2022.11.14.22282195

**Authors:** Jay A. Pandit, Jennifer M. Radin, Danielle Chiang, Emily Spencer, Jeff Pawelek, Mira Diwan, Leila Roumani, Michael Mina

**Author notes:** **Corresponding author:** Jay A. Pandit, MD, Scripps Research Translational Institute, 3344 N Torrey Pines Ct. Plaza Level, La Jolla, CA 92037, USA.

## Abstract

**Introduction:** The uptake of Paxlovid in individuals infected with COVID-19 has been significantly limited by concerns around the Paxlovid rebound phenomenon despite the scarcity of evidence around its epidemiology. The purpose of this study was to prospectively compare the epidemiology of Paxlovid rebound in treated and untreated participants with acute COVID-19 infection

**Methods:** We designed a decentralized, digital, prospective observational study in which participants who tested positive for COVID-19 using eMed Test-to-Treat telehealth kits and were clinically eligible for Paxlovid were recruited to be evaluated for viral and symptom clearance, as well as rebound. Participants were assigned to a Paxlovid or control group based on their decision to take Paxlovid. Following initial diagnosis based on a telehealth proctored test both groups were provided 12 telehealth proctored rapid antigen home tests and asked to test on a regular frequent schedule for 16 days and answer symptom surveys. Viral rebound based on test results and COVID-19 symptom rebound based on patient reported symptoms were evaluated.

**Results:** Viral rebound incidence was 14.2% in the Paxlovid group (n=127) and 9.3% in the control group (n=43). COVID-19 symptom rebound incidence was higher in the Paxlovid group (18.9%) compared to the control group (7.0%). There were no notable differences in viral rebound by age, gender, pre-existing conditions, or major symptom groups during the acute phase or at the 1-month interval.

**Conclusion:** This preliminary report of our prospective study suggests that rebound after clearance of test positivity or symptom resolution is higher than previously reported. However, we observed a similar rate of rebound in both in the Paxlovid and control groups. Large studies with diverse participants and extended follow-up are needed to better understand the rebound phenomena.

## Introduction

COVID-19 continues to be a global health crisis that is responsible for over 6 million reported deaths so far [1]. Oral antiviral medications such as Paxlovid and Molnupravir are two of the main pharmaceutical interventions for preventing progression of symptoms in non-hospitalized patients [2, 3]. The EPIC-HR (Evaluation of Protease Inhibition for Covid-19 High-Risk Patients) study showed that patients treated with Paxlovid had an 89% lower risk to develop severe symptoms, compared to placebo [4]. As the first oral antiviral agent on the market, the government committed to purchasing 10 million Paxlovid treatment doses, followed by another tranche to support test to treatment programs during the omicron wave [5]. However, Paxlovid remains largely under-prescribed due to concerns around medication interactions and the peculiar Paxlovid rebound phenomenon.

With increasing use of Paxlovid, several publications reported a return of COVID-19 symptoms or detectable viral load, after completion of the Paxlovid course [6-9]. This “rebound” phenomenon occurs in a subset of COVID-19 patients who have completed the 5-day course of Paxlovid, subsequently testing negative for SARS-CoV-2 virus, and two to eight days later experience a temporary return of symptoms with or without a positive rapid antigen or real-time polymerase chain reaction (RT-PCR) test [10]. Further viral rebound and symptom rebound have also been seen in participants who did not receive any treatment leading to viral or symptom resolution [11], imploring further investigation into this rebound phenomenon. There have been some suggestions that rebound could be related to drug pharmacodynamics [12] or anti-viral mediated interactions with viral immunologic response [13], however, peer reviewed literature on Paxlovid rebound is mostly limited to retrospective case series with wide estimates [7, 9, 14-16]. Rebound in untreated cohorts has consisted of small prospective studies [11, 17].

Gaining a better understanding of demographic and clinical characteristics associated with developing rebound is important for future tailoring treatment length, timing of treatment, or advising certain populations who may be at greater risk of developing rebound. Rebound may result in unintended transmission after someone has tested negative and return of symptoms may discourage people from getting treatment to prevent severe disease progression. The purpose of this study is to prospectively compare the epidemiology of Paxlovid rebound in participants with acute COVID-19 infection who receive Paxlovid as compared to Paxlovid eligible controls who independently choose to not receive Paxlovid. In this manuscript we present the first report of our ongoing study which will continue to enroll participants and track them over a longer period.

## Methods

### Study Design and Recruitment

We designed a decentralized, digital, prospective observational study developed through a collaboration between Scripps Research Translational Institute (SRTI) and eMed (a virtual care platform that assists users through at-home testing, and automatically triages them to a telemedicine visit where they may be prescribed treatment, if eligible). For COVID-19, this process is called eMed digital “Test-to-Treat.” Participants qualified for the study if they were 18 years of age or older, had a positive rapid antigen test for SARS-CoV-2 verified by eMed Telehealth proctoring (standard in the Test-to-Treat process), and were prescribed Paxlovid through the eMed telehealth (regardless of whether they intended to take it). After being recorded as COVID-19 positive and clinically evaluated, participants who were offered Paxlovid then received an email within hours with a link to the study materials, including informed consent. Participants who chose to participate were shipped a study kit overnight as described below. Participants who resided outside of the U.S. and/or did not speak English were excluded.

### Study Procedures and Assessments

Interested participants were split into two arms based on their independent decision to take Paxlovid. All study tasks were identical in both arms (Paxlovid and control; Figure 1). After the informed consent process, participants completed a baseline demographic and pre-existing conditions survey. Enrolled participants were shipped, overnight, a kit with twelve eMed telehealth proctored Abbott BinaxNOW COVID-19 rapid antigen home tests. Participants in the Paxlovid arm completed their first study-provided antigen test and their first symptom surveys on days 2 and 5 of the 5-day Paxlovid course (day 2 was the quickest the study kits could be delivered to participants), and then every other day through day 16. After the 16-day period, participants completed a persistent/long COVID symptoms survey at 1-, 3-, and 6-month intervals.

**Figure 1:**
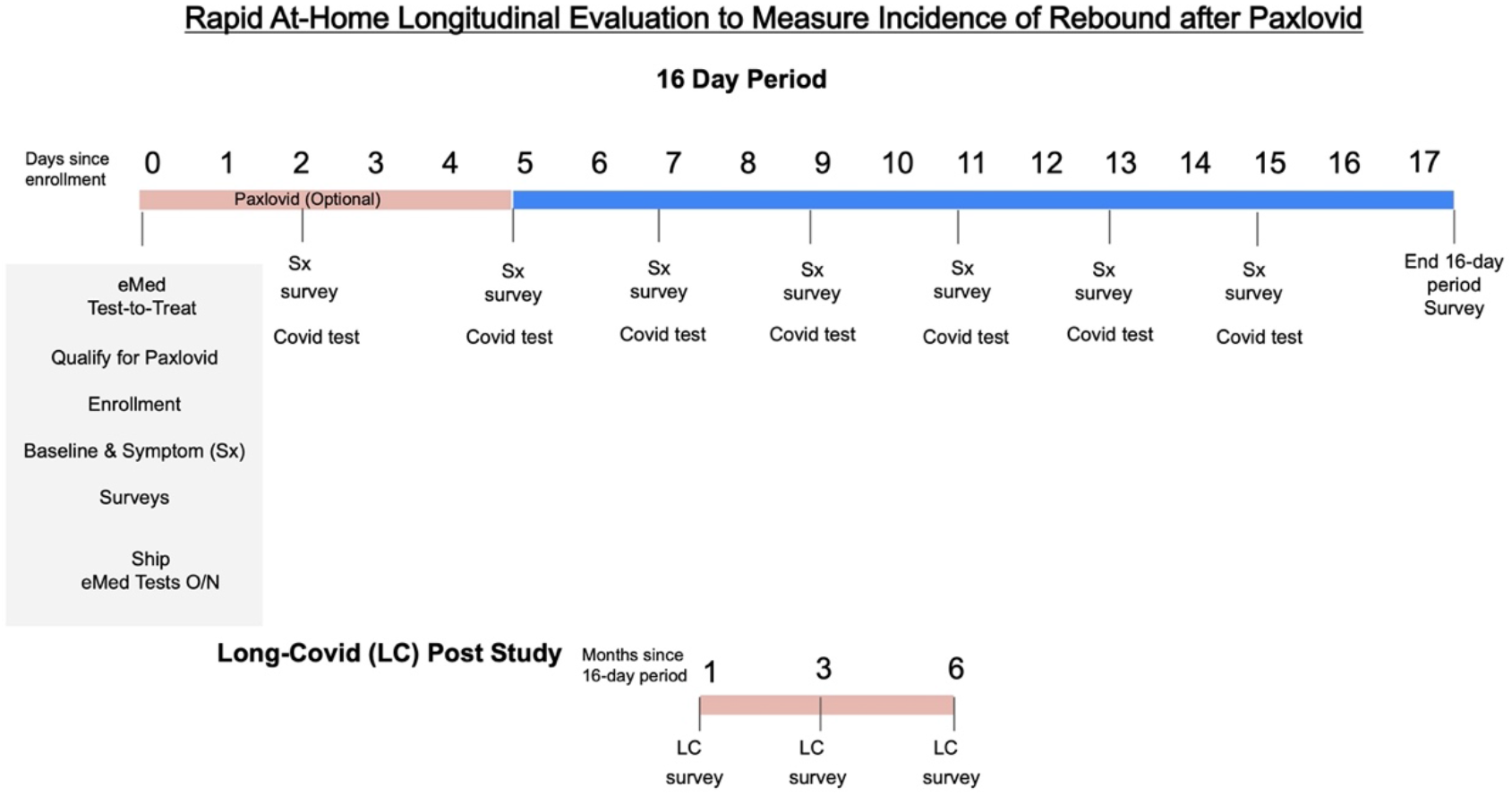
Study Tasks: After consent, participants were asked to take a rapid antigen COVID-19 test and a symptom survey on day 2, day 5, day 7, day 9, day 11, day 13 and day 15. Participants were then given an end of 16-day period survey, followed by a long covid symptom survey at the 1-, 3-, and 6-month periods.

### Definitions

#### Viral (Testing) rebound

Any participant with a positive rapid antigen test observed after a negative antigen test.

#### Non-rebound for Viral (Testing)

Any participant with a positive test, followed by only negative tests within the 16-day (every other day testing) study period.

#### Symptom rebound

Any participant who reported resolution of symptoms and then a recurrence on subsequent symptom surveys within the acute 16-day study period.

#### Symptoms definitions

Respiratory (cough, runny nose, shortness of breath or difficulty breathing, sore throat, hoarse voice), Gastrointestinal (nausea, vomiting, diarrhea, stomachache, loss of appetite), Neurologic (headache, confusion) and Systemic (fevers, chills, shaking with chills, loss of taste/smell, muscle pain, chest pain, eye pain, body ache, fatigue, neck pain, rash).

### End Points

The primary endpoint was the incidence of viral (testing) and symptom rebound within the Paxlovid and control groups after acute COVID-19 infection. Secondary endpoints were time to test negativity, time to symptom resolution, time from negative test to rebound positive test and frequency of symptoms in both groups during the acute phase and the 1-month mark.

### Oversight

The study was conducted in accordance with Good Clinical Practice Guidelines and was approved by the Scripps Institutional Review Board. Electronic informed consent was obtained from all the participants. The study was designed in collaboration with the sponsor (eMed). Safety oversight was performed by Scripps. Study data were collected and managed using Research Electronic Data Capture (REDCap) tools hosted at SRTI [18, 19]. REDCap is a secure, web-based software platform designed to support data capture for research studies, providing 1) an intuitive interface for validated data capture; 2) audit trails for tracking data manipulation and export procedures; 3) automated export procedures for seamless data downloads to common statistical packages; and 4) procedures for data integration and interoperability with external sources.

### Statistical Analysis

Kaplan-Meier survival curves, stratified by Paxlovid arm, were plotted to compare time to events in each arm. Participants were censored at the time of two missing tests or missing symptom surveys in a row (for test-related results or symptom-related results, respectively), or by day 15 (last test day) if they did not have an event. A log-rank test was used to compare the survival curves of the treatment and control groups. Participant characteristics and COVID-19 recovery categories were compared (using a Chi-squared test for categorical and t-test for continuous variables) between the two groups as well as differences between individuals who did and did not rebound, regardless of treatment group and reported symptoms at month 1 between the treatment and control group (Table 3). All analyses were done in SAS version 9.4. Of note, because this the first report of our ongoing study, the current report is not fully powered to evaluate statistical significance. Thus, we do not attempt to draw statistical conclusions of group differences but focus, rather, on reporting observations and incidence.

## Results

### Participants

Between August 4, 2022 and November 1, 2022, 247 participants consented to participate in the study, and 188 participants had completed the 16-day study procedures. Participants with less than two completed surveys or test results or who completed the surveys outside the outlined survey schedule were excluded from the analysis, leaving 170 participants: 127 participants in the Paxlovid treatment arm and 43 in the control arm for the analysis.

The treatment and control arms had no notable differences by age and gender, or pre-existing conditions (Table 1). There was a higher frequency of white participants in the Paxlovid group compared to controls (p=0.0002). Five individuals (4%) in the Paxlovid group remained positive throughout the 16-day period, and thus were not eligible for evaluation for rebound and were excluded from the tables but were included in the time to negative test and symptom resolution analyses.

**Table 1.** Participant characteristics and COVID-19 recovery by Paxlovid treatment, frequency (%)

|  | Paxlovid<br>(n=127) | No Paxlovid (n=43) | p-value |
| --- | --- | --- | --- |
| Participant Characteristics |  |  |  |
| Race/ethnicity |  |  |  |
| White | 111 (88.1) | 26 (61.9) | 0.0002 |
| African American | 9 (7.1) | 8 (19.1) | 0.03 |
| Asian | 5 (4.0) | 3 (7.1) | 0.40 |
| Latino | 10 (7.9) | 6 (14.3) | 0.22 |
| Hawaiian/Pacific<br>Islander | 1 (0.8) | 0 (0.0) | 0.56 |
| American Indian | 5 (4.0) | 3 (7.1) | 0.40 |
| Age Category (years old) |  |  | 0.42 |
| 18-44 | 51 (40.2) | 22 (51.2) |  |
| 45-64 | 58 (45.7) | 17 (39.5) |  |
| 65+ | 18 (14.2) | 4 (9.3) |  |
| Gender |  |  | 0.48 |
| Female | 72 (56.7) | 27 (62.8) |  |
| Male | 55 (43.3) | 16 (37.2) |  |
| Pre-existing Conditions |  |  |  |
| Asthma | 21 (16.5) | 8 (18.6) | 0.76 |
| Cancer | 6 (4.7) | 1 (2.3) | 0.49 |
| Heart Disease | 3 (2.4) | 2 (4.7) | 0.44 |
| Heart Failure | 0 (0.0) | 1 (2.3) | 0.08 |
| COPD | 0 (0.0) | 1 (2.3) | 0.08 |
| High Blood Pressure | 34 (26.8) | 12 (27.9) | 0.88 |
| Chronic Bronchitis | 1 (0.8) | 1 (2.3) | 0.42 |
| Other lung condition | 2 (1.6) | 0 (0.0) | 0.41 |
| Diabetes | 10 (7.9) | 2 (4.7) | 0.48 |
| Autoimmune<br>Condition | 12 (9.5) | 1 (2.3) | 0.13 |
| Emphysema | 0 (0.0) | 1 (2.3) | 0.08 |
| None of above | 56 (44.1) | 25 (58.1) | 0.11 |
| Received at least 1 Covid-19 Vaccine Dose | 121 (95.3) | 41 (95.3) | 0.98 |
| COVID-19 Recovery |  |  |  |
| Viral (Testing) Rebound | 18 (14.2) | 4 (9.3) | 0.41 |
| Symptom Rebound | 24 (18.9) | 3 (7.0) | 0.06 |
| Consistently Positive | 5 (3.9) | 0 (0.0) | 0.20 |
| Symptom Start to Test<br>Negative, Days (SD)* | 6.4 (3.0) | 6.1 (2.9) | 0.53 |
| Symptom Start to First Day of No Symptoms, Days (SD)* | 8.4 (3.7) | 8.5 (4.3) | 0.90 |
| First Positive Test to First Negative Test, Days (SD)* | 6.8 (2.9) | 7.0 (3.9) | 0.73 |

\*Only includes participants who had an event from Figure 2 (Tested Negative or Reported No Symptoms).

Time to viral clearance, defined as time from first positive antigen test to first negative antigen test was similar in the treatment and control groups (mean 6.8 days vs. 6.7 days). Similarly, time from symptom onset to first symptom resolution (mean 10.4 days vs.10.7 days), and time from symptom onset to first negative antigen test (mean 6.0 days vs. 6.3 days) was similar in the treatment and control group (Figure 2 and Table 1). However, the numbers above only included people who had an event (tested negative or symptoms resolved) during the 15-day time period. Many participants were still positive and/or symptomatic at day 15 which makes the actual time an underestimate. Approximately 20% of individuals in both groups were still positive on a rapid antigen test at 10 days after first turning antigen test positive (Figure 2a).

**Figure 2.**
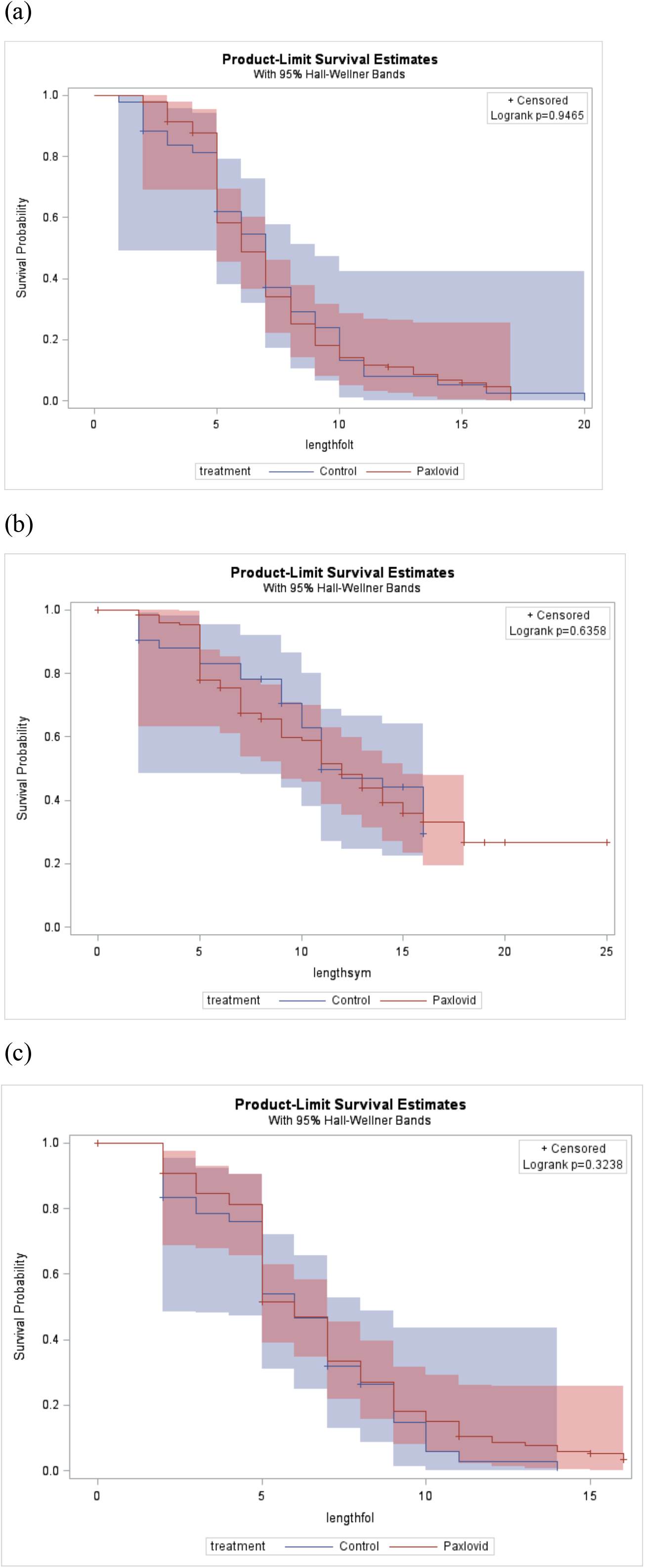
Survival curves comparing Paxlovid treatment and control groups. Comparisons of (a) first positive test to first antigen negative test; (b) symptom start to first report of no symptoms; (c) symptom start to first antigen negative test. (a)

### Rebound Incidence

Virus testing rebound incidence trended higher in the Paxlovid group (18/127; 14.2%) than in the control group (4/43; 9.3%) (Figure 3a and Table 1). Symptom rebound incidence was notably higher in the Paxlovid group (18.9%) than the control group (7.0%). Participants who rebounded also reported less body ache compared to those who did not rebound. There were no notable differences in viral rebound by age, gender, pre-existing conditions, or symptom groups during the acute 16-day follow-up period. When comparing participants with viral rebound versus those who did not have viral rebound, frequency of Asian and Native American participants was lower among those with rebound (Table 2). Further, no notable differences were observed in the treatment and control group symptoms at the 1-month period. (Table 3)

**Table 2.** Participant characteristics by viral rebound (includes Paxlovid treatment and control groups), frequency (%).

|  | Rebound (n=22) | No Rebound (n=148) | p-value |
| --- | --- | --- | --- |
| Race/ethnicity |  |  |  |
| White | 15 (71.4) | 122 (83.0) | 0.20 |
| African American | 2 (9.5) | 15 (10.2) | 0.92 |
| Asian | 3 (14.3) | 5 (3.4) | 0.03 |
| Latino | 2 (9.5) | 14 (9.5) | 1.00 |
| Hawaiian/Pacific Islander | 0 (0.0) | 1 (0.7) | 0.70 |
| American Indian | 3 (14.3) | 5 (3.4) | 0.03 |
| Age Category (years old) |  |  | 0.09 |
| 18-44 | 9 (40.9) | 64 (43.2) |  |
| 45-64 | 7 (31.8) | 68 (46.0) |  |
| 65+ | 6 (27.3) | 16 (10.8) |  |
| Gender |  |  | 0.19 |
| Female | 10 (45.5) | 89 (60.1) |  |
| Male | 12 (54.6) | 59 (39.9) |  |
| Pre-existing Conditions |  |  |  |
| Asthma | 2 (9.1) | 27 (18.2) | 0.29 |
| Cancer | 2 (9.1) | 5 (3.4) | 0.21 |
| Heart Disease | 0 (0.0) | 5 (3.4) | 0.38 |
| Heart Failure | 0 (0.0) | 1 (0.7) | 0.70 |
| COPD | 0 (0.0) | 1 (0.7) | 0.70 |
| High Blood Pressure | 7 (31.8) | 39 (26.4) | 0.59 |
| Chronic Bronchitis | 0 (0.0) | 2 (1.4) | 0.58 |
| Other lung condition | 1 (4.6) | 1 (0.7) | 0.12 |
| Diabetes | 1 (4.6) | 11 (7.4) | 0.62 |
| Autoimmune Condition | 1 (4.6) | 12 (8.1) | 0.56 |
| Emphysema | 0 (0.0) | 1 (0.7) | 0.70 |
| None of above | 10 (45.5) | 71 (48.0) | 0.83 |
| Symptoms |  |  |  |
| Body Ache | 6 (27.3) | 86 (58.1) | 0.007 |
| Symptom Groups |  |  |  |
| Systemic | 15 (68.2) | 124 (83.8) | 0.08 |
| Neurologic | 12 (54.6) | 98 (66.2) | 0.29 |
| Gastrointestinal | 6 (27.3) | 46 (31.1) | 0.72 |
| Respiratory | 22 (100.0) | 146 (98.7) | 0.58 |
| Received at least 1 COVID-19 Vaccine Dose | 22 (100.0) | 140 (94.6) | 0.26 |

**Table 3.** Symptoms reported at month 1 between Paxlovid treatment and control groups, frequency (%).

|  | Paxlovid (n=109) | No Paxlovid (n=34) | p-value |
| --- | --- | --- | --- |
| Palpitations | 3 (2.8) | 0 (0.0) | 0.33 |
| Neurological | 5 (4.6) | 0 (0.0) | 0.20 |
| Sleep Disturbance | 2 (5.9) | 2 (1.8) | 0.21 |
| Mental | 3 (2.8) | 0 (0.0) | 0.33 |
| Menstrual | 1 (0.9) | 0 (0.0) | 0.58 |
| Headache | 10 (9.2) | 2 (5.8) | 0.55 |
| Body aches | 9 (8.3) | 4 (11.8) | 0.53 |
| Cough | 19 (17.4) | 7 (20.6) | 0.68 |
| Congestion | 11 (10.1) | 6 (17.7) | 0.23 |
| Sore Throat | 4 (3.7) | 2 (5.9) | 0.57 |
| Headache | 10 (9.2) | 2 (5.9) | 0.55 |
| Fatigue | 19 (17.4) | 8 (23.5) | 0.43 |
| SOB | 9 (8.3) | 1 (2.9) | 0.29 |
| Tinnitus | 1 (0.9) | 1 (2.9) | 0.38 |

**Figure 3.**
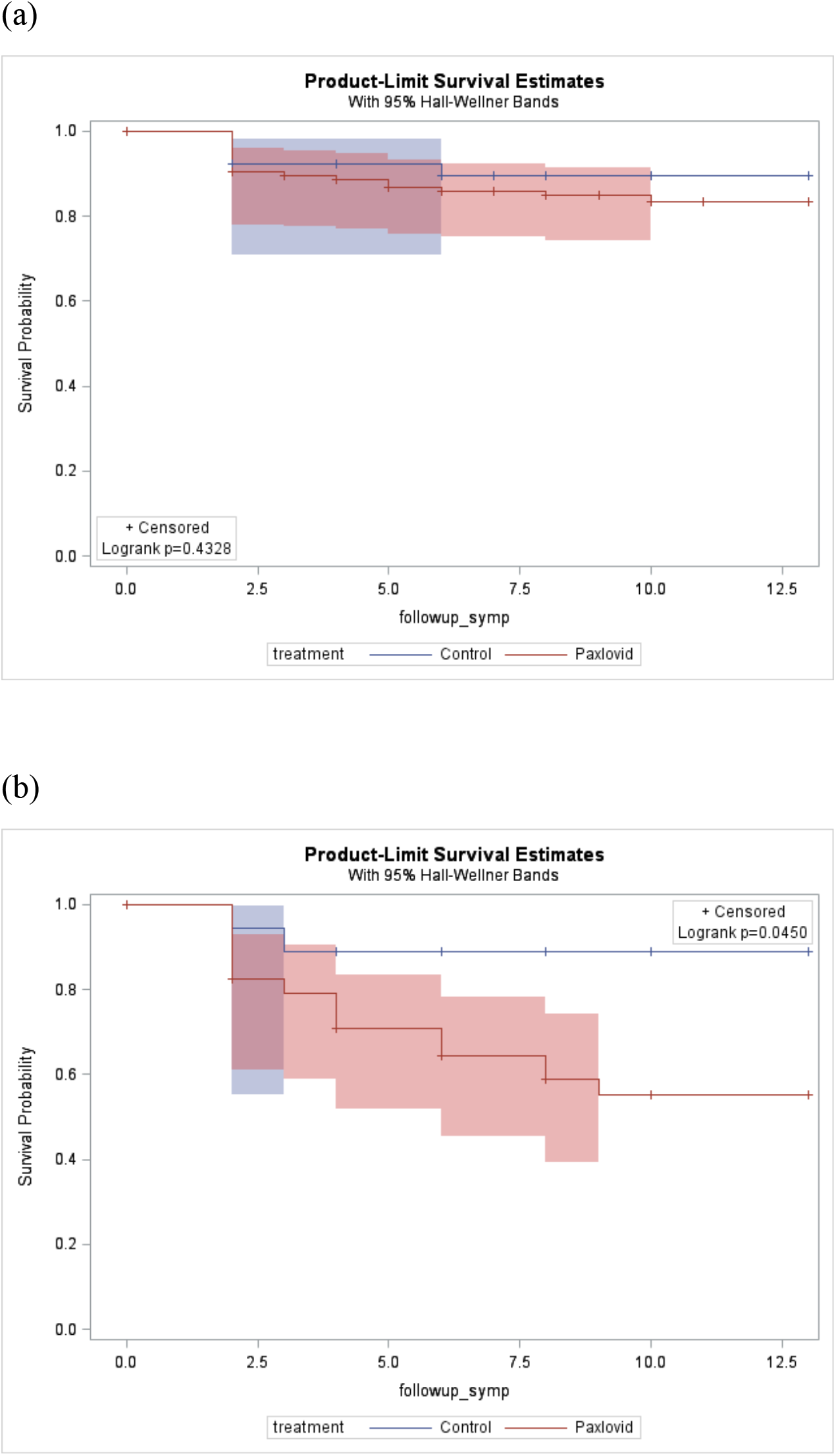
Survival curves between Paxlovid treatment and control groups (a). Time from negative test to viral rebound (testing positive) grouped by Paxlovid treatment. Only includes individuals with event (tested negative) from figure 2a. (b) Time from no symptoms to symptom rebound grouped by Paxlovid treatment. Only includes individuals with event (reported no symptoms) from figure 2b.

Out of the control group, 2 had a symptom rebound that lasted less than 5 days and 1 had a rebound that was 5 days or longer. In the Paxlovid group 10 had a symptom rebound that was less than 5 days, 10 had symptom rebound that was 5 days or more and 4 had more than 1 symptom rebound during the 16-day follow-up period.

## Discussion

The current indication for Paxlovid is for the treatment of mild-to-moderate COVID-19 in adults with positive SARS-CoV-2 viral testing and who are at a high risk for progression to severe COVID-19, including hospitalization or death, yet there is no consensus on what to do when Paxlovid or untreated individuals rebound. Our study is one of the first to prospectively evaluate Paxlovid rebound with a study population that is acutely positive with COVID-19 and comparable in age and gender to the EPIC-HR study [4]. In addition to the study results reported, this study demonstrates the feasibility of offering a test to treatment approach, collecting patient reported outcomes and clinical outcomes in a decentralized fashion, which is ideal for participants acutely infected with COVID-19 and complements public health prevention measures.

Our study demonstrated an overall viral testing rebound incidence of 14% and a symptom rebound incidence of 19% among Paxlovid treated cases. Both incidences are higher than have been reported in prior retrospective studies which ranged from 2% to 6% [20, 21]. However, we also show that both viral (9%) and symptom rebound (7%) occurs in the absence of treatment with Paxlovid. Testing rebound in the control group matches reported viral rebound incidence in other studies of untreated patients with COVID-19, which was 12% [11, 17]. A notable finding in our study was that symptom rebound in the control cohort was lower than in the Paxlovid arm. Though the small sample size limits our ability to draw statistical inference, this finding could be driven by the pharmacology of Paxlovid. The current predominant hypothesis around rebound is that the immune system may not have the opportunity or need to fully ramp up upon infection with the virus, since Paxlovid suppressed replication early in disease [10]. However, in our cohort, Paxlovid treatment was not associated with a shorter time to symptom resolution or testing negative. It is possible that there is not an increase in viral rebound after taking Paxlovid but rather more rebound is identified in this group due to heightened awareness of the possibility, and studies have supported that in both treatment and control groups the rebound symptoms are milder than symptoms of the COVID-19 infection [22]. While Paxlovid treatment reduces severe outcomes [4], the high incidence of viral and symptom rebound in both treated and untreated cohorts suggests that investigation is needed around the changes caused by Paxlovid in the virologic and immunologic milieu of the pathogen and host.

We identified several interesting signals (acknowledging the study was not powered for statistical inference) in participant characteristics and acute phase symptoms among the treatment and control group. We saw a significantly higher Paxlovid uptake among white participants and less among black participants, which has also been noted by the CDC [20]. Understanding barriers to use is important for improving COVID-19 outcomes. We also saw a significantly lower frequency of Asian Americans experiencing viral rebound and less rebound in individuals who didn’t report body aches during the acute phase. In the future, identifying risk factors for developing rebound could help tailor treatment duration in specific subpopulations and help inform participants to retest if they are in a high rebound risk group.

Positive rapid antigen tests are associated with levels of virus that remain infectious. In this study we observed that over 50% of participants remained positive on a rapid antigen test at five days after first turning positive or becoming symptomatic, and 20% of participants remained positive even ten days into infection. Thus, our study agrees with others that have found that the 5-day isolation period, as currently recommended by the CDC, is not adequate. Rather than using a single cut-off for how long to remain in isolation, a “test to exit” approach would enable individuals to take a data driven approach to their decision making around assuming they are no longer infectious and a risk to those around them.

For our next phase, we will collect testing swabs for viral sequencing as well as serum from participants to understand any virus specific or host specific factors that provide insight into Paxlovid rebound. Retrospective analyses have reported the lack of an association between viral load rebound and low nirmatrelvir exposure or resistance to nirmatrelvir, however, they were done during the B.1.617.2 (delta) variant rather than the B1.1.529 (omicron) variant [23]. Notably, the EPIC-HR study focused on viral load based rebound only, which does not translate into the presence of infectious virus, and more importantly does not include symptom rebound [4]. The strengths of our study include its prospective data collection, during the predominant omicron wave and decentralized manner, which makes it pragmatic.

The primary limitations of the study were the unbalanced sample size in the control cohort and the largely white population in the cohort, which will be actively addressed in the follow up study by using a target percentage for underrepresented populations in biomedical research. The primary goal of the current study was to understand the incidence of Paxlovid rebound and therefore we accepted the cohorts in a convenience sampling approach. Other sources of bias include recruitment through the eMed platform, which could introduce a selection bias to eMed customers. Further, participants who consented to participate were sent the research kit and were told to start testing right after they started Paxlovid, however, some participants waited a few days to begin testing or started Paxlovid after testing, which we attempted to adjudicate as best as possible. Additionally, to balance participation burden with adherence, we asked participants to do every other day testing and did not ask participants to do daily testing and surveys, which could lead to missing data points.

This study adds vital incidence information for Paxlovid rebound and spotlights the need to better understand the rebound phenomenon of our main pharmaceutical therapeutics options for the COVID-19 pandemic response.

## Conclusion

This preliminary report of our prospective study of both virus test-positive and symptomatic rebound suggests that rebound after clearance of test positivity or symptom resolution is higher than previously reported. Interestingly, however, we observed that the high rate of rebound is both in the Paxlovid treatment and control groups. Large studies with diverse participants and extended follow-up are needed to better understand the rebound phenomena.

## Data Availability

This is an ongoing study. We plan to make the de-identified data available after approval of a proposal by a responsible authority at Scripps and with a data access agreement, pledging to not re-identify individuals or share the data with a third party.

